# Barriers to Clinical Trial Efficiency and Patient Access: The Heart Failure Collaboratory Industry Survey

**DOI:** 10.1101/2020.06.24.20124917

**Authors:** Shashank S. Sinha, Mitchell A. Psotka, Mona Fiuzat, Scott D. Barnett, Martina Bruckmann, Javed Butler, Mary M. DeSouza, G. Michael Felker, Scott D. Solomon, Norman Stockbridge, John R. Teerlink, Ellis F. Unger, Christopher M. O’Connor, Marvin A. Konstam

**Author notes:** Correspondence to: Marvin A Konstam, MD, FACC, Chief Physician Executive, The CardioVascular Center, Tufts Medical Center, Box 108, 800 Washington Street, Boston, MA 02111.

## Abstract

**Aims:** Clinical trial inefficiency and lack of patient access to novel therapies have been identified as key barriers to successful heart failure innovation. The Heart Failure Collaboratory (HFC) is a consortium developed to identify and address barriers to bringing drugs and devices to market. The HFC performed an electronic cross-sectional survey of key leaders within industry to collate and interpret a sample of viewpoints on these challenges.

**Methods and Results:** From August to September 2018, self-administered survey data were electronically collected from industry partners. Group comparisons were made via Fisher’s Exact or chi-square test. Respondents most commonly rated the United States Food and Drug Administration (44.2%), Health Canada (39.5%) and European Medicines Agency (32.6%) as efficient. Respondents rated the top 3 areas with the greatest opportunity for regulatory agencies to improve efficiency: ‘improve usefulness of agency feedback’ (48.8%); ‘improved timeliness of agency responses’ (41.9%); and ‘pre-specification of required magnitude of clinical effect’ and ‘limit excessive data requirements’ (32.6%). Respondents rated items of ‘excessive clinical site staff workload’ (55.8%), ‘overly complex case report forms’ (51.2%) and ‘data input errors’ (39.5%) as the top 3 factors influencing data quality. Respondents rated items of ‘onerous prior authorization requirements’ (51.2%), ‘availability of decision-making rationale’ (46.5%) and ‘rationality of access barriers created by payers’ (44.2%) as the top 3 impediments to improving patient access to approved therapies.

**Conclusion:** This broadly distributed survey of industry respondents identified multiple specific barriers to heart failure clinical trial efficiency and patient access, which should help direct efforts toward improvement.

## INTRODUCTION

A number of factors have been identified as posing challenges to heart failure drug and device development: limited patient engagement in clinical research, an erratic culture of participation, slow evidence generation, a declining clinical research infrastructure, complicated study design, high costs, diminishing return on investment, and access barriers following regulatory approval.^1-6^ The Heart Failure Collaboratory (HFC) is a consortium of heart failure research stakeholders including patients, investigators, academic leaders, pharmaceutical and device industry representatives, medical society representatives, third-party payers, and government representatives from the United States Food and Drug Administration (FDA), National Institutes of Health (NIH), and Centers for Medicare and Medicaid Services (CMS). Founded in March 2017, the purpose of the HFC is to identify barriers to bringing effective drugs and devices to patients and to seek ways to mitigate those barriers (www.hfcollaboratory.com).

There are at least 3 broad categories of obstacles to the availability of new therapies to patients: the regulatory approval process; clinical trial performance; and establishment of payer coverage, payment, and access.^7,8^ To assess industry perceptions of the relative impact of specific factors, across these categories, in hindering the delivery of new drugs and devices to patients, the HFC performed an electronic cross-sectional survey of industry sponsors active in heart failure therapy development. We analyzed the responses to gain insight toward prioritizing efforts to accelerate the rates at which heart failure therapies reach patients.

## METHODS

### Subjects and Settings

This cross-sectional survey of industry sponsors active in the field of developing novel drug and device therapies for heart failure was designed to describe and rank perceived barriers to treatment delivery. Participants included industry medical executives who self-identified as developing therapies for heart failure and for whom contact information was available.

From August to September 2018, self-administered survey data were electronically collected from industry partners. The survey was first distributed to the entire HFC mailing list and associated colleagues by electronic mail after a pre-notification mailing was sent using the Qualtrics platform. All HFC members were potential participants. Responses were monitored closely, and reminders were distributed to the target population three times before the process closed. Additional promotion encouraging survey participation was conducted through electronic and verbal communications at the Heart Failure Society of America’s 22^nd^ Annual Scientific Meeting in September 2018, and through face to face meetings of the HFC.

### Survey Instrument

The HFC Drugs and Devices Working Groups designed the electronic survey to describe the involvement of industry sponsors in clinical trial data collection and record the perceived challenges impairing clinical trial efficiency and post-regulatory availability of therapies. Potential barriers and demographic variables that may modify those barriers were initially identified by review of the published literature. Stakeholders from the HFC Working Groups were included in the subsequent semi-structured focused discussions regarding item generation, reduction, and pre-testing. The survey was pilot tested within the HFC Working Groups to ensure appropriate content and to improve readability and clarity prior to full distribution (see Supplementary Appendix for full content).

The resulting HF Collaboratory Industry Survey was a 76-item survey comprising 3 domains: a) *Issues Related to the Regulatory Process; b) Issues Related to Trial Conduct*; and c) *Issues Related to Coverage and Payment*, with 10 sub-domains ranging from *Improving Efficiency* to *Improving Patient Access*. The scale response utilized two Likert-type rating scales, of either adversely influencing efficiency (1 = major influence; 5 = minimal influence) or improvement opportunity (1 = the least opportunity; 6 = the most opportunity).

### Statistical Analysis

Data are presented as means ± standard deviation or frequency and percent. Group comparisons were made via Fisher’s exact or chi-square test, where appropriate. All statistical analyses were conducted using SAS (ver. 9.3, Cary NC). Two-sided p-values ≤ 0.05 were considered statistically significant, without correction for multiple comparisons. The survey was deemed eligible for exemption under 45 CFR 46.101(b)(2) according to the Office for Human Research Protections (OHRP).

## RESULTS

One hundred and twenty-five participants were emailed a link to initiate and complete the online survey. Seventy-five participants initiated the online questionnaire with 43 participants (‘*survey completers*’) completing the survey for an overall response rate of 34.4% (Figure 1). These 43 completers constituted the analysis cohort. The characteristics of the survey group are described in Table 1.

**Table 1.** Characteristics of Survey Respondents (n=75)

| Parameter | Survey<br>Non-Completer<br>N=32 | Survey<br>Completer<br>N=43 | Total<br>N=75 |
| --- | --- | --- | --- |
| Which category best describes your experience with industry? |  |  |  |
| Clinical Development/R&D | 20 (62.5) | 25 (58.1) | 45 (60.0) |
| Clinical Operations | 12 (37.5) | 18 (41.9) | 30 (40.0) |
| Are you a member of: |  |  |  |
| Global team only | 13 (40.6) | 22 (51.2) | 35 (47.7) |
| US and Global team | 19 (59.4) | 21 (48.8) | 40 (53.3) |
| Which of the following best describes your company's products? |  |  |  |
| Drugs | 19 (59.4) | 20 (46.5) | 39 (52.0) |
| Device | 8 (25.0) | 8 (18.6) | 16 (21.3) |
| Drugs and Device | 5 (15.6) | 15 (34.9) | 20 (26.7) |

R&D, research and development; US, United States

**Figure 1.**
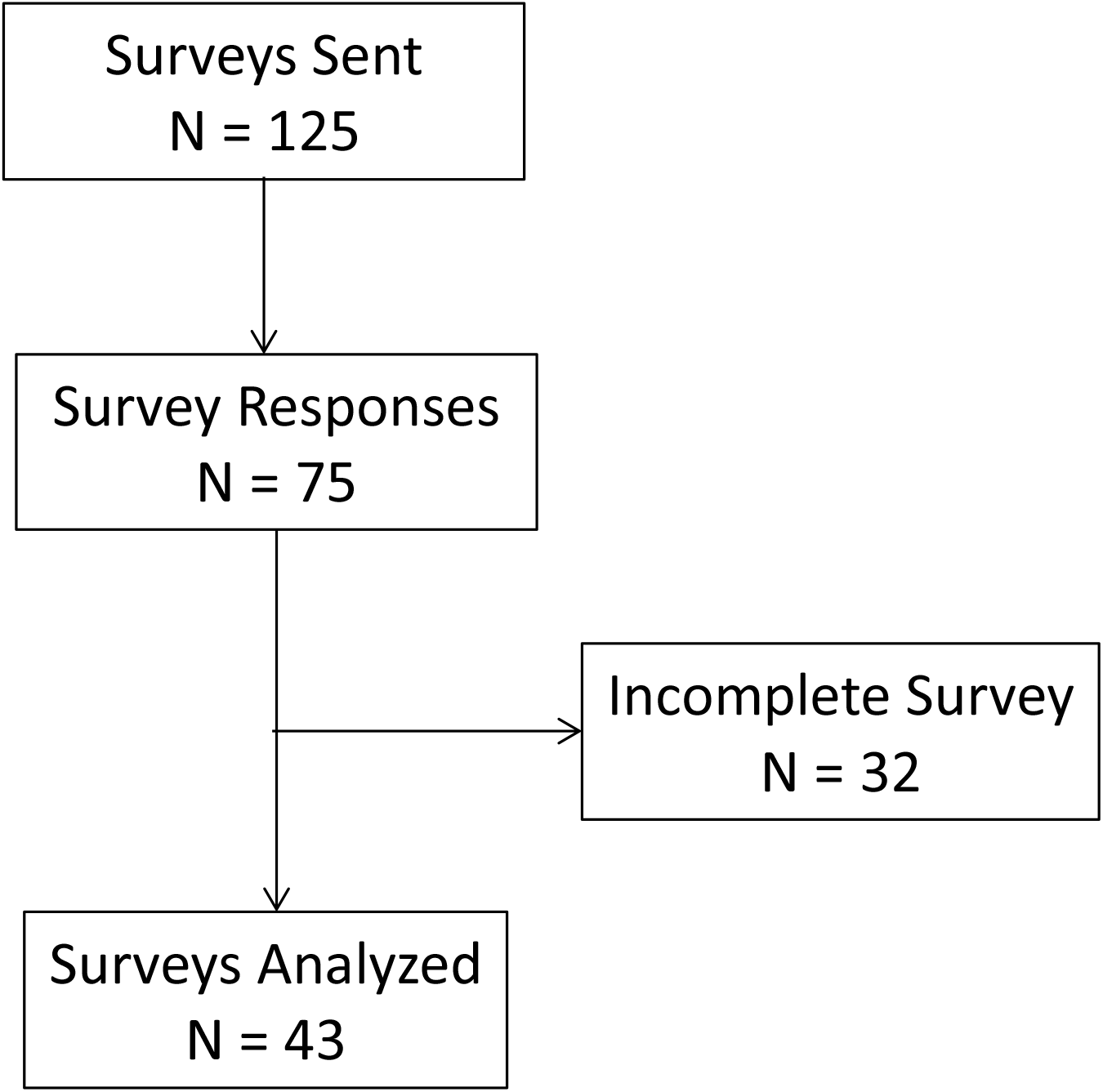
Flow Diagram for Inclusion and Exclusion of Survey Respondentsx

## ISSUES RELATED TO THE REGULATORY PROCESS

### Regulatory Efficiency

Respondents most commonly rated the United States Food and Drug Administration (FDA) (44.2%), Health Canada (39.5%) and the European Medicines Agency (EMA) (32.6%) as efficient (Figure 2A). Respondents rated the top 3 areas with the greatest opportunity for regulatory agencies to improve their efficiency as: ‘*improved usefulness of agency feedback’* (48.8%); ‘*improved timeliness of agency responses*’ (41.9%); and ‘*pre-specification of required magnitude of clinical effect*’ (32.6%) and ‘*limit excessive data requirements*’ (32.6%) (Figure 2B). Areas rated as less important are shown in Supplemental Table 1.

**Figure 2.**
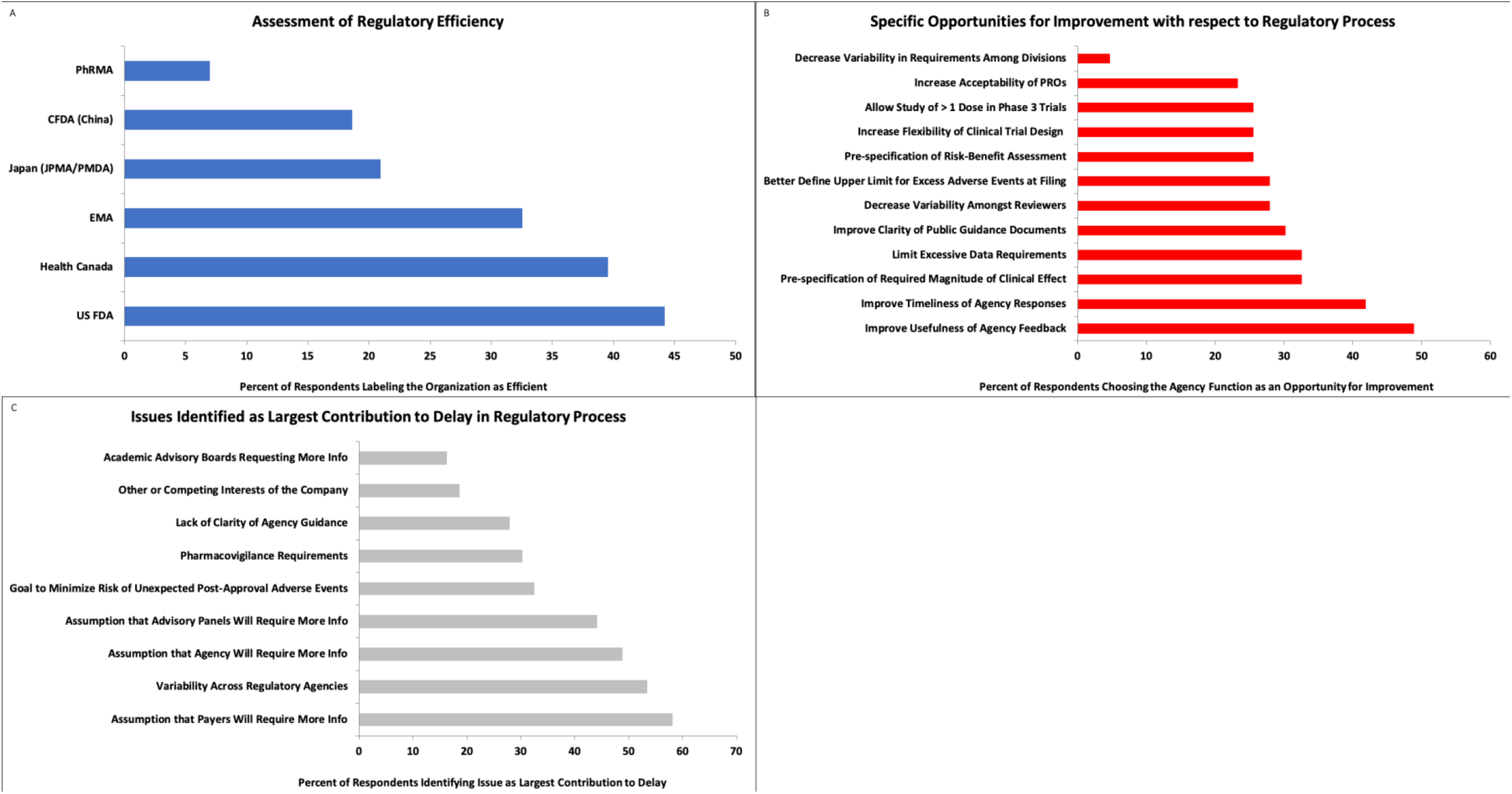
Respondent Rated Items of Issues Related to the Regulatory Process. Figure 2A) Assessment of Regulatory Efficiency. Figure 2B) Specific Opportunities for Improvement with respect to the Regulatory Process. Figure 2C) Issues Identified as Largest Contribution to Delay in Regulatory Process. Abbreviations. CFDA; EMA = European Medicines Agency; FDA = Food and Drug Administration; JPMA/PMDA = Japan Pharmaceuticals and Medical Devices Agency; PhRMA = Pharmaceutical Research and Manufacturers of America; PROs = patient-reported outcomes.

### Data Collection

When participants were asked to identify the largest contributors to regulatory delay phase 3 clinical trials, participants rated the top 3 reasons as ‘*assumption that payers will require more information’* (58.1%), ‘*variability across regulatory agencies*’ (53.5%), and ‘*assumption that agency will require more information*’ (48.8%) (Figure 2C). Other factors are shown in Supplemental Table 1.

## ISSUES RELATED TO TRIAL CONDUCT

### Study initiation

Respondents rated ‘*budget negotiations and contracting*’ (51.2%), ‘*insufficient resources at clinical sites*’ (39.5%) and ‘*vendor contracting*’ (34.9%) as the largest impediments to study initiation (Figure 3A) Other factors are listed in Supplemental Table 2.

**Figure 3.**
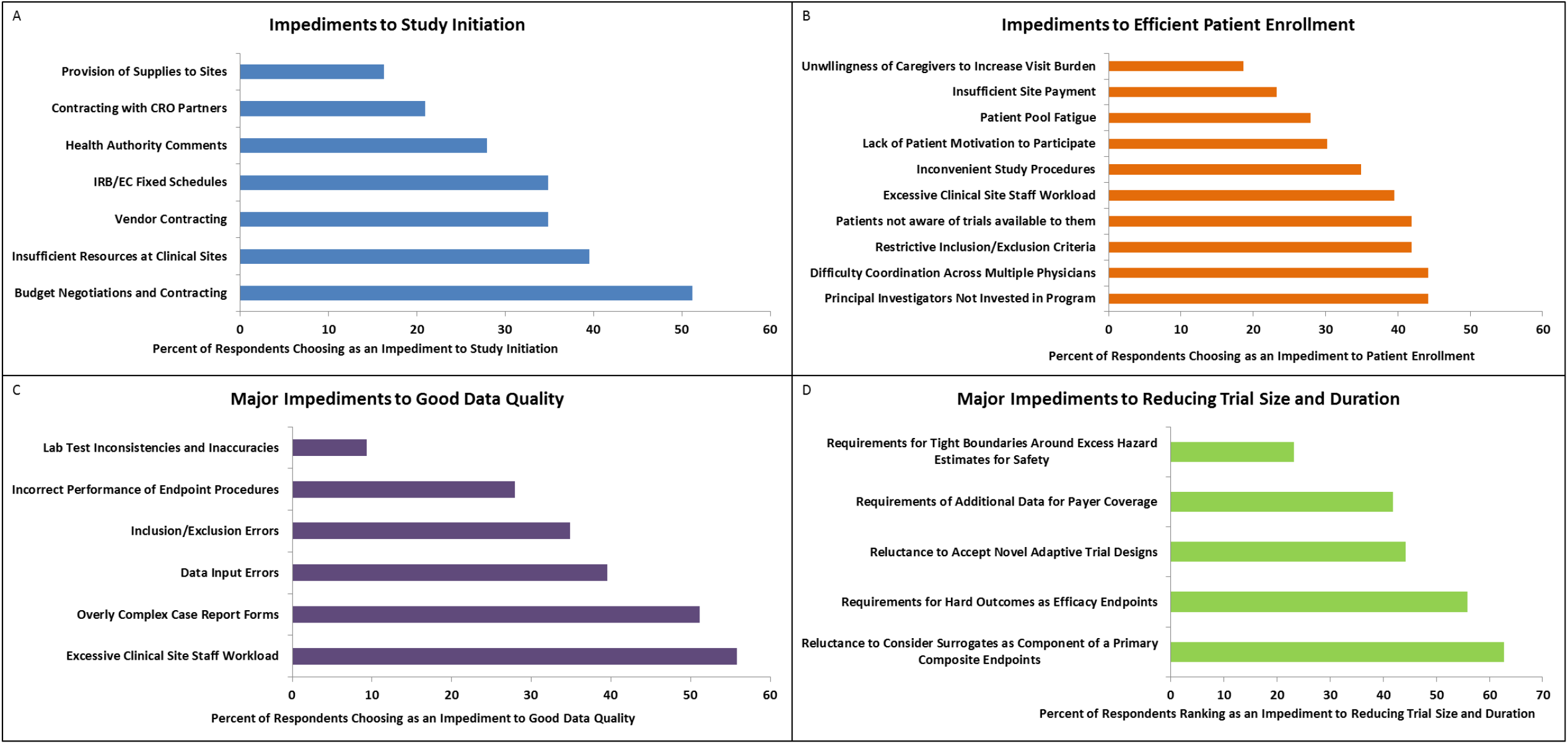
Respondent Rated Items of Issues Related to the Trial Conduct. Figure 3A) Impediments to Study Initiation. Figure 3B) Impediments to Efficient Patient Enrollment. Figure 3C) Major Impediments to Good Data Quality. Figure 3D) Major Impediments to Reducing Trial Size and Duration. Abbreviations: CRO = contract research organization; EC = ethics committee; IRB = institutional review board.

### Patient Enrollment

Respondents rated items of ‘*principal investigator not invested*’ (44.2%), ‘difficulty in *coordination across multiple physicians*’ (44.2%), and ‘*restrictive inclusion/exclusion criteria*’ (41.9%) as the top 3 impediments to patient enrollment (Figure 3B). Other factors are listed in Supplemental Table 2.

### Data Quality

Respondents rated items of ‘*excessive clinical site staff workload*’ (55.8%), ‘overly complex *case report forms (CRF)*’ (51.2%) and ‘*data input errors*’ (39.5%) as the top 3 factors influencing data quality (Figure 3). Other factors are shown in Supplemental Table 2.

### Reducing Trial Size and Duration

Respondents rated items of ‘*reluctance to consider surrogates as component of primary composite endpoints*’ (62.8%), ‘*requirement for hard outcomes as efficacy endpoints*’ (55.8%) and ‘*reluctance to accept novel adaptive trial design*’ (44.2%) as the top 3 factors impeding reduction of trial sample size (Figure 3D). Other factors are shown in Supplemental Table 2.

## ISSUES RELATED TO COVERAGE, PAYMENT, AND ACCESS

### Obtaining Access to Markets

Respondents rated ‘*insufficient evidence to obtain high valuation score*’ (51.2%), ‘*requirements for active comparators in trials versus use of real world evidence (RWE)*’ (46.5%) and ‘*consolidation of United States market and access to formularies*’ (37.2%) as the top 3 impediments to obtaining market access (Figure 4A). Other factors are shown in Supplemental Table 3.

**Figure 4.**
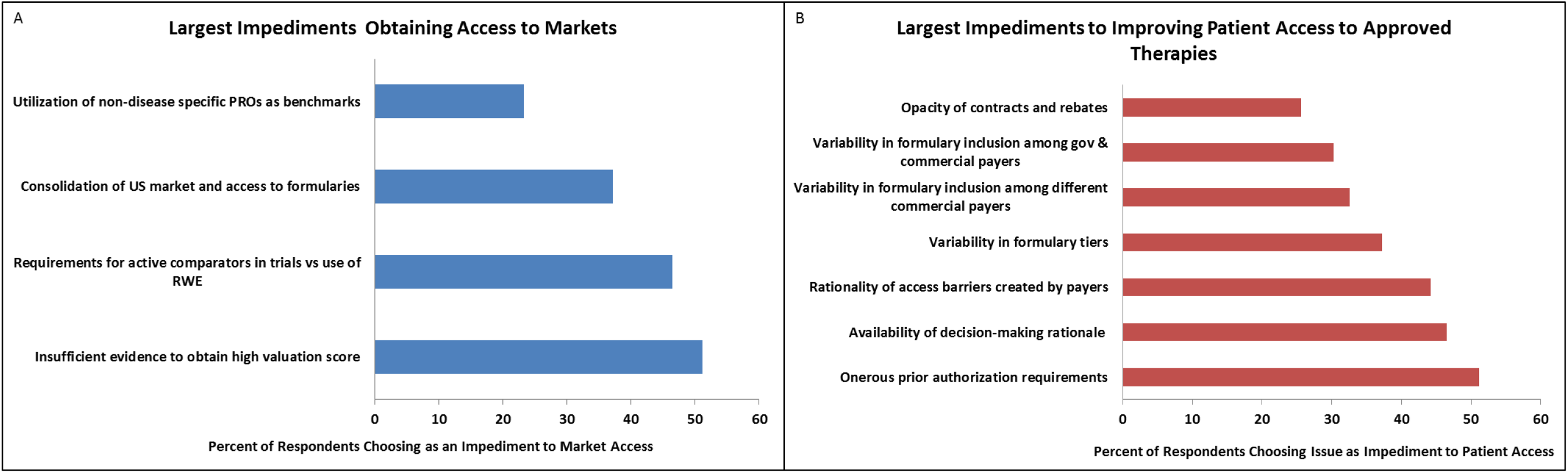
Respondent Rated Items of Issues Related to Coverage, Payment, and Access. Figure 4A) Largest Impediments to Obtaining Access to Markets. Figure 4B) Largest Impediments to Improving Patient Access to Approved Therapies. Abbreviations: gov = government; PROs = patient reported outcomes; RWE = real-world evidence; US = United States.

### Improving Patient Access to Approved Therapies

Respondents rated items of ‘*onerous prior authorization requirements*’ (51.2%), ‘*availability of decision-making rationale*’ (46.5%) and ‘*rationality of access barriers created by payers*’ (44.2%) as the top 3 impediments to improving patient access to approved therapies (Figure 4B). Other factors are shown in Supplemental Table 3.

## DISCUSSION

The HFC Working Group designed the electronic survey to rank the perceptions of industry representatives in relation to challenges in regulatory efficiency, clinical trial design, clinical trial efficiency, and patient access to novel therapies. The findings of this broadly distributed survey suggest numerous barriers exist to delivering novel, safe, and effective treatments to patients with heart failure.

Over-collection of data was commonly cited as a cause for regulator inefficiency. The top reasons respondents reported over-collection of data were concern that more data would be required – either by regulatory agencies or by payers – and variability in requirements across regulatory agencies. Over-collection leads to increased complexity, cost, and duration of clinical trials. These findings highlight the importance of clarifying essential data needs and to harmonize requirements, both across regulators from different world regions, and between regulators and payers. Efforts are now underway to achieve greater harmonization between the US FDA and EMA with respect to data standards for efficacy and safety.^9^ The FDA has issued guidance regarding the extent of safety data collection in late-stage premarket and post-approval clinical trials.^10^

Clinical trial performance was another area identified by respondents with significant opportunity for improvement. The principal culprit impairing data quality was identified to be the manner in which data are collected. Specifically cited were the workload of clinical site staff and complexity of CRFs, likely related and contributing to data entry errors and protocol violations.^8^ The HFC has expended considerable effort to define a standardized minimal data set CRF, with participants from academia, industry, and regulatory agencies, as well as patients.^11^ These instruments may be supplemented by modules, where additional types of data, such as echocardiographic measurements are defined in a standardized manner. Another potential advance is to move rapidly toward electronic data entry across all trials and sites. In fact, one might envision a day when electronic medical records might be so standardized, powerful, and connected that most data required for trials could be extracted directly from medical records, obviating the need for tedious manual data entry and reducing the impact of human error.^12,13^

Patient access to approved drugs and devices presents a myriad of challenges. Industry responses in this area highlight a decision-making system that is perceived to be 1) onerous, 2) arbitrary, 3) opaque, and 4) focused more on restricted utilization than on following the scientific evidence to drive patient benefit.^14-17^ Here, opinions from industry representatives appear to be in harmony with those from clinicians and patients in asking for multiple improvements in the processes and decision-making on insurance coverage. The HFC has called for substantially increased transparency and rationality in regard to middle-man contracts, rebates, and the legitimacy of rules driving prior authorization processes.

The HFC is poised to utilize the findings of this survey to foster solutions and initiatives among clinicians, industry partners, regulators, and payers.^2^ A new generation of researchers provides opportunities for novel research design, novel trial endpoints, and the innovative use of comparator groups. These new methodological approaches to clinical trials may use existing foundational research paradigms to augment current drug or device development and simultaneously provide growth in clinical research design. Indeed, the HFC is engaged in several initiatives to address these issues, including the establishment of standardized heart failure trial definitions and endpoints, trial design, data collection, and statistical analytic methods, encouragement processes for high-performing research networks, and strategies to ease access to novel therapies.

Limitations to this survey include a modest 34% response plus completion rate despite multiple attempts to enhance engagement. In fact, this rate is substantially higher than that seen for many surveys, though a higher rate would have rendered our conclusions even more credible and generalizable. Further potential sources of error include unrecognized systematic biases that may be present in the survey questionnaire, or potentially constituencies of drug and device development that may have been under-represented, such as developers of biologic compounds and diagnostic tests.

In summary, results of this survey provide insight into the directions that should be prioritized in accelerating and improving the regulatory process and clinical trial performance, and reforming the structures behind insurer decision-making. This effort illustrates the potential for active and open partnerships across academia, industry, regulators, payers and patients toward developing and approving novel therapies more rapidly and providing greater access to those therapies, based on solid clinical trial evidence.

## Data Availability

The primary and corresponding author take full responsibility for primary data collection and analysis.

## Acknowledgements

We would like to gratefully acknowledge the thoughtful contributions and participation of the HFC drug working group members, including but not limited to Chris Cabell, Graziella Collu, Peter DiBattiste, Joerg Koglin, Marty Lefkowitz, Lothar Roessig, and Steven Simonson.

---

**Supplemental Table 1.** Issues Related to the Regulatory Process (n=43).

| Item | N (%) |
| --- | --- |
| Agencies maximizing efficiencies as '*Best*' |  |
| US FDA | 19 (44.2) |
| Health Canada | 17 (39.5) |
| EMA | 14 (32.6) |
| Japan (JPMA/PMDA) | 9 (20.9) |
| CFDA (China) | 8 (18.6) |
| PhRMA | 3 (7.0) |
| Agency's opportunities for improved efficiency as '*Most Efficient*' |  |
| US FDA | 20 (46.5) |
| EMA | 9 (20.9) |
| Health Canada | 2 (4.7) |
| Japan (JPMA/PMDA) | 1 (2.3) |
| CFDA (China) | 0 (0.0) |
| PhRMA | 0 (0.0) |
| Magnitude of Opportunity for Improved Efficiency as '*Most Opportunity*' |  |
| Usefulness / clarity of agency feedback / guidance in response to industry queries and meetings. | 21 (48.8) |
| Timeliness of agency responsiveness | 18 (41.9) |
| An excessive amount of observational data required for collection pre-NDA | 14 (32.6) |
| Pre-specification of clinically acceptable efficacy magnitude | 14 (32.6) |
| Clarity and extent of public guidance documentation | 13 (30.2) |
| Variability across reviewers within the same division | 12 (27.9) |
| Upper boundary for excess morbidity / mortality at time of filing of an NDA/BLA | 12 (27.9) |
| Pre-specification of guidance regarding quantifiable risk-benefit profile | 11 (25.6) |
| Flexibility of design (e.g. adaptability) | 11 (25.6) |
| Barriers to studying more than one dose in phase 3 | 11 (25.6) |
| Greater acceptability of patient reported outcomes | 10 (23.3) |
| Variability in requirements across different divisions | 2 (4.7) |
| A problem for contributing to the amount of data required by the agency during phase 3 for approvability as the '*Largest Contributor*' |  |
| Assumption that more information is going to be required later, by payers | 25 (58.1) |
| Variability across global regulatory agencies | 23 (53.5) |
| Assumption that more information is going to be required later by the agency | 21 (48.8) |
| Assumption that more information is going to be requested later by advisory panels | 19 (44.2) |
| Industry desire to minimize the risk of unexpected adverse events arising post-approval | 14 (32.6) |
| Pharmacovigilance requirements within the company | 13 (30.2) |
| Lack of clarity of agency guidance | 12 (27.9) |
| Other interests of the company beyond the current specific application | 8 (18.6) |
| Academic advisory boards requesting information for future publications | 7 (16.3) |

\*Agencies are sorted highest to lowest

**Supplemental Table 2.** Issues Related to the Trial Conduct.

| Item | N (%) |
| --- | --- |
| Terms as the '*Largest Impediment*' to efficient study initiation |  |
| Budget Negotiations/Contracting with Clinical Study Sites/PIs | 22 (51.2) |
| Insufficient/inadequate resources at clinical sites | 17 (39.5) |
| Vendor contracting and/or reconciliation of vendor SOPs with CRO/Sponsor SOPs | 15 (34.9) |
| IRB/ EC/ fixed schedule/ comments/ delays to approval | 15 (34.9) |
| Health Authority extensive comments/delays/ approvals | 12 (27.9) |
| Contracting with CRO partners | 9 (20.9) |
| Delays in provision of study/clinical supplies to clinical sites | 7 (16.3) |
| Factors that may influence patient enrollment as the '*Largest Impediment*' |  |
| Coordination of multiple physicians/departments in order to qualify patients for study. | 19 (44.2) |
| Clinical PI not invested in program/lack of motivation | 19 (44.2) |
| Excessively restrictive study inclusion/exclusion criteria | 18 (41.9) |
| Patients not aware of trials available to them | 18 (41.9) |
| Excessive clinical site staff workload | 17 (39.5) |
| Study procedures are inconvenient for trial staff or patient (late nights, weekends. | 15 (34.9) |
| Patients not motivated to participate in trials | 13 (30.2) |
| Same sites as prior, patient pool fatigue | 12 (27.9) |
| Insufficient/low site payment | 10 (23.3) |
| Caregivers are not willing to take patients to multiple visits | 8 (18.6) |
| Factors that may influence data quality as the '*Largest Impediment*' |  |
| Excessive clinical site staff workload | 24 (55.8) |
| CRF complexity; difficulty entering data | 22 (51.2) |
| Deficiencies in site training; data input errors | 17 (39.5) |
| Clinical site not trained properly, patient inclusion/exclusion errors | 15 (34.9) |
| Endpoint procedures (e.g. PROs, 6MWT, ECHO) not performed correctly | 12 (27.9) |
| Local lab/central lab test inconsistency / inaccuracy | 4 (9.3) |
| Perceived impediments to reducing necessary trial size as the '*Largest Impediment*' |  |
| Reluctance to consider validated surrogate endpoints as components of a primary endpoint composite | 27 (62.8) |
| Requirement for hard outcomes, as opposed to patient reported outcomes, as (effectively) the only acceptable efficacy endpoints for registration | 24 (55.8) |
| Reluctance to accept novel adaptive trial designs | 19 (44.2) |
| Requirements of additional data for payer coverage | 18 (41.9) |
| Requirement for tight boundaries around excess hazard estimates for safety endpoint | 10 (23.3) |

**Supplemental Table 3.** Issues Related to Coverage, Payment, and Access.

| Item | N (%) |
| --- | --- |
| Perceived impediments to obtaining access to markets as the '*Largest impediment*' |  |
| Sufficient evidence to obtain high valuation score requirement (Europe) | 22 (51.2) |
| Need for active comparator in trial versus use of real world registry observational data as comparator | 20 (46.5) |
| Continual consolidation of US market and access to formulary (e.g. Insurer-PBM consolidation, Health Institution/Insurer consolidation) | 16 (37.2) |
| Utilization of non-disease specific PROs (EQ-5D) as benchmarking with regard to access and pricing | 10 (23.3) |
| Areas of improving patient access to approved therapies as the '*Greatest opportunity*' |  |
| Excessively onerous prior authorization requirements and processes | 22 (51.2) |
| Availability of decision-making rationale used by governmental and commercial payers/insurers regarding coverage & cost-sharing decisions | 20 (46.5) |
| Rationality of access barriers created by payers/insurers | 19 (44.2) |
| Variability in formulary tiers, and coinsurance/copayment requirements from payer to payer | 16 (37.2) |
| Variability in formulary inclusion between different commercial payers/insurers | 14 (32.6) |
| Variability in formulary inclusion between governmental and commercial | 13 (30.2) |
| Opacity of contracts and rebates between insurers, pharmacies, pharmacy benefit managers that determine other accessibility decisions | 11 (25.6) |

